# Modulation of a Stable Neurobehavioral Trait Using Repetitive Transcranial Magnetic Stimulation: A Preregistered Randomized Controlled Trial

**DOI:** 10.1101/2021.07.08.21260222

**Authors:** Afik Faerman, James H. Bishop, Katy H. Stimpson, Angela Phillips, Merve Gülser, Heer Amin, Romina Nejad, Danielle D. DeSouza, Andrew D. Geoly, Elisa Kallioniemi, Booil Jo, Nolan R. Williams, David Spiegel

## Abstract

Hypnotizability, one’s ability to experience cognitive, emotional, behavioral, and physical changes in response to suggestions in the context of hypnosis, is a highly stable trait associated with increased functional connectivity between the left dorsolateral prefrontal cortex (L-DLPFC) and dorsal anterior cingulate cortex (dACC). We conducted a preregistered, triple-blinded, randomized controlled trial to test the ability of continuous theta-burst stimulation (cTBS) over a personalized neuroimaging-based L-DLPFC target to temporarily enhance hypnotizability. We tested our hypothesis in 78 patients with fibromyalgia syndrome (FMS), a functional pain disorder for which hypnosis has consistently been shown to be beneficial as a nonpharmacological treatment option. Pre-to-post cTBS change in Hypnotic Induction Profile scores (HIP; a standardized measure of hypnotizability) was significantly greater in the Active versus Sham group. Our findings suggest a causal relationship between L-DLPFC and dACC function and hypnotizability. Dose-response optimization should be further examined to formalize guidelines for future clinical utilization.

**Trial registration:** ClinicalTrials.gov NCT02969707

## INTRODUCTION

Hypnosis was the first Western conception of psychotherapy^1^. Hypnosis can facilitate the treatment and management of numerous psychiatric and neurological symptoms^2^, yet not all people respond to hypnosis equally. Hypnotizability, an individual’s capacity to respond to suggestions given in hypnosis, is a trait comprised of cognitive, neural, and behavioral components,^3,4^ and has been demonstrated to moderate the effects of hypnosis-based interventions^5^. Approximately two-thirds of the general adult population are hypnotizable, and 15% are highly hypnotizble^6^. Hypnotizability has been shown to be a surprisingly stable trait within individuals throughout adulthood, with .7 test-retest correlations over a 25 year interval^7^. Although stable, the modulation of trait hypnotizability has been investigated for decades. Gorassini and Spanos^8,9^ developed in-person sessions based on behavioral training, resulting in increased hypnotic responsiveness. Despite its relative success in modifying participants’ responsiveness in hypnosis, not all replication studies yielded similar effects^10,11^, and behavioral training has failed to elicit increases in responsiveness in large numbers of individuals^12^. Lynn and colleagues argued that inherent neurocognitive differences between “naturally” high-hypnotizable and low-hypnotizable individuals might explain the limits of a behavioral intervention in modifying hypnotizability^13^. Several clinical trials attempted modifying hypnotic responsiveness using psychoactive drugs and other chemical substances, including Lysergic Acid Diethylamide (LSD-25), mescaline, psilocybin^14^, diazepam^15^, nitrous oxide^16^, oxytocin^17^, and modafinil^18^. However, such substances have substantial effects on perception and behavior and different pharmacological pathways pose conceptual limitations on interpretation and replication of hypnotizability enhancement, and potential risks limit the practicality of their use in clinical settings. Further randomized clinical trials are needed to evaluate these issues and explore emerging pathways to uncover modifiable hypnotizability characteristics. Localized, nonpharmacological neuromodulation offers a more accurate route to test local neurobiological mechanisms with relatively fewer risks and precise replicability. It provides the opportunity to test the possibility of altering a highly stable neurobiological trait, allowing for causal as well as correlational inference.

Neurocognitively, hypnotizability is associated with functional connectivity between the left dorsolateral prefrontal cortex (L-DLPFC) and the dorsal anterior cingulate cortex (dACC)^4^, two central nodes for executive control and conflict processing, respectively. Although the activity of specific brain regions during hypnosis is largely task-dependent high hypnotizability is associated with altered activations of the anterior cingulate and prefrontal cortices^19,4^. A recent study by our group extended on this finding by showing that the levels of the inhibitory neurotransmitter GABA in the anterior cingulate cortex (ACC) were positively associated with higher hypnotizability^20^. Structural findings further suggest that greater gray matter volume in the medial frontal cortex and ACC positively correlate with increased hypnotic depth ratings^21^. Together, these data suggest a circuit underlying trait hypnotizability and provide a potential target for noninvasive neuromodulation.

Transcranial magnetic stimulation (TMS) noninvasively facilitates neuronal interactions using a high-intensity magnetic field induced brief, focal electric field in the cortex. This electric field is able to activate neurons. Repetitive TMS (rTMS) produces periods of lasting facilitation or inhibition that persist after stimulation^22^. Similar to hypnosis, low frequency rTMS application to the DLPFC is associated with decreased DLPFC activity and increased functional connectivity with the dACC^23,24^. Due to the shared neural effects of hypnotizability and rTMS application to the DLPFC, it is reasonable to infer that modulating DLPFC-ACC interactions with rTMS may result in temporary changes in hypnotizability. Indeed, a previous study showed that inhibitory rTMS, applied to the left DLPFC, increased both objective and subjective experiences of hypnotic responsiveness in a small cohort of medium hypnotizables^25^.

A briefly administered and clinically accessible intervention to temporarily increase hypnotizability might improve the effectiveness of hypnosis-based treatments and their relevance to various populations. This is a particularly salient clinical opportunity in populations where the first line of treatment is limited in effectiveness or safety, and hypnosis has been demonstrated as a valid treatment alternative. In this study, we focused on providing causal evidence for the neural mechanism of trait hypnotizability and the possibility of modulating it via neurostimulation in a clinical population. Thus, we conducted a triple-blind, randomized controlled study to determine whether we can utilize rTMS to modulate hypnotizability in patients with fibromyalgia syndrome (FMS), a functional pain disorder for which hypnosis has consistently been shown to be beneficial as a nonpharmacological treatment option^26^. We hypothesized that inhibitory rTMS application to the L-DLPFC would significantly increase hypnotizability compared to sham stimulation.

## METHODS

### Participants

Eighty-one low to moderately hypnotizable male and female participants with FMS aged 18-69 years were recruited for this study starting in February 2017, with both recruitment and data collection concluding due to targeted enrollment number being reached in December 2019. The study was approved by the Stanford University Institutional Review Board (IRB), and all participants provided informed consent. Prior to enrollment, participants underwent both phone and in-person screening procedures to determine eligibility. Hypnotizability was assessed during the in-person screening by trained study personnel using the Hypnotic Induction Profile (HIP; see below). Low to moderately hypnotizable individuals (≤8/10 HIP score) were eligible to participate in the study, and highly hypnotizable individuals (>8/10 HIP score) were excluded. In addition to meeting the hypnotizability requirements, all participants had a primary diagnosis of FMS, which was confirmed by a study clinician during the in-person screening. Diagnostic criteria were determined based on the American College of Rheumatology Preliminary Diagnostic Criteria for fibromyalgia syndrome^27^, and participants provided a blood sample to confirm normal complete blood count (CBC) and inflammatory panel. Exclusionary criteria included standard magnetic resonance imaging (MRI) contraindications (e.g., ferromagnetic implants, claustrophobia), neurological disorders (e.g., seizure disorder), and psychiatric disorders (e.g., bipolar disorder, schizophrenia). Given the high comorbidity rate of depression and chronic pain syndromes, particularly fibromyalgia^28^, depressive symptoms were not exclusionary. However, participants with severe Major Depressive Disorder or depressive symptoms with suicidal ideation were not enrolled in the study. Participants with fibromyalgia currently prescribed psychoactive medications underwent a voluntary washout period prior to neuroimaging and transcranial magnetic stimulation that was individually tailored to the participant by a study psychiatrist. To assure blinding, only participants with no previous exposure to TMS were eligible for the study. See Figure 1 for a recruitment Consort diagram.

**Figure 1.**
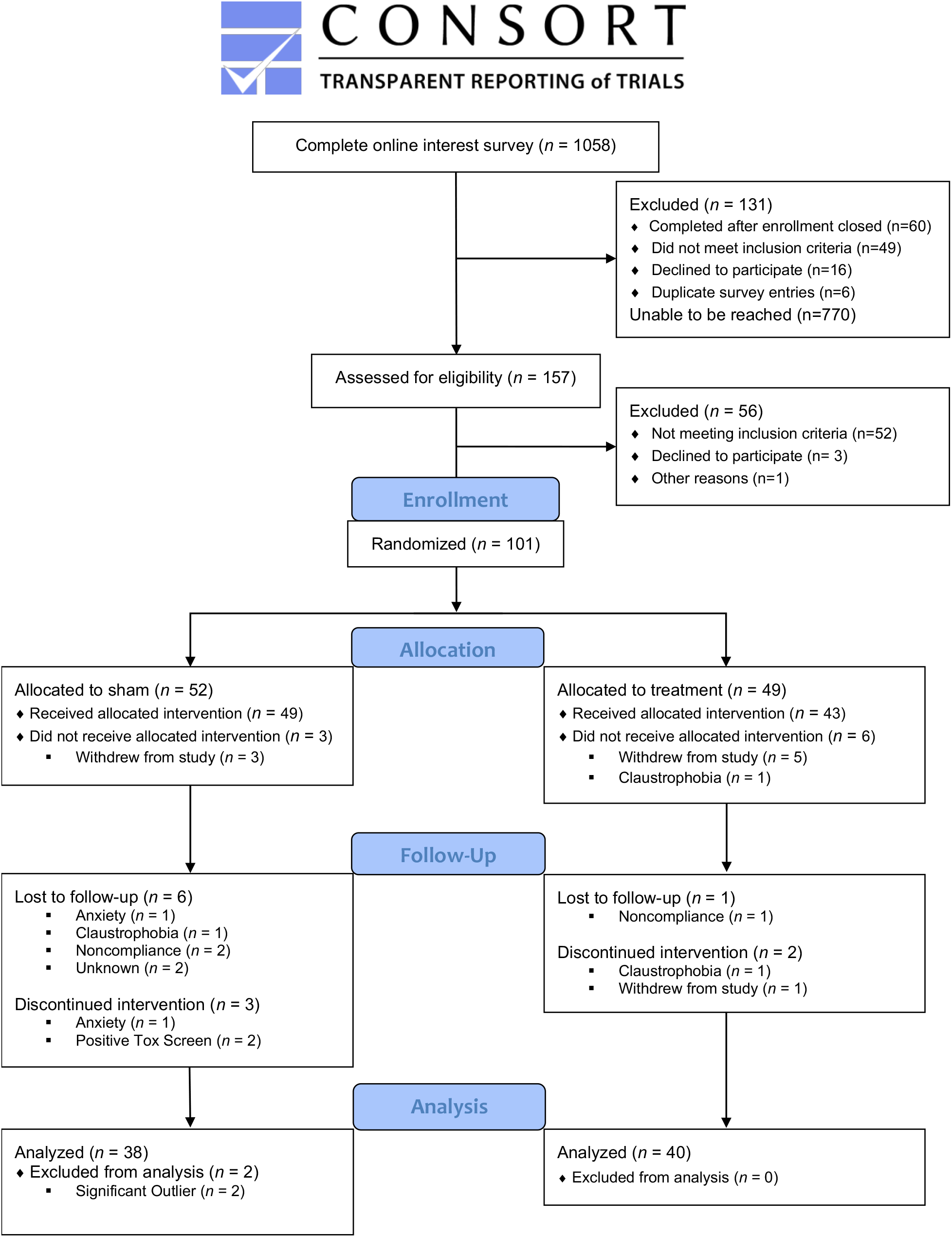
Recruitment Consort diagram.

**Figure 2.**
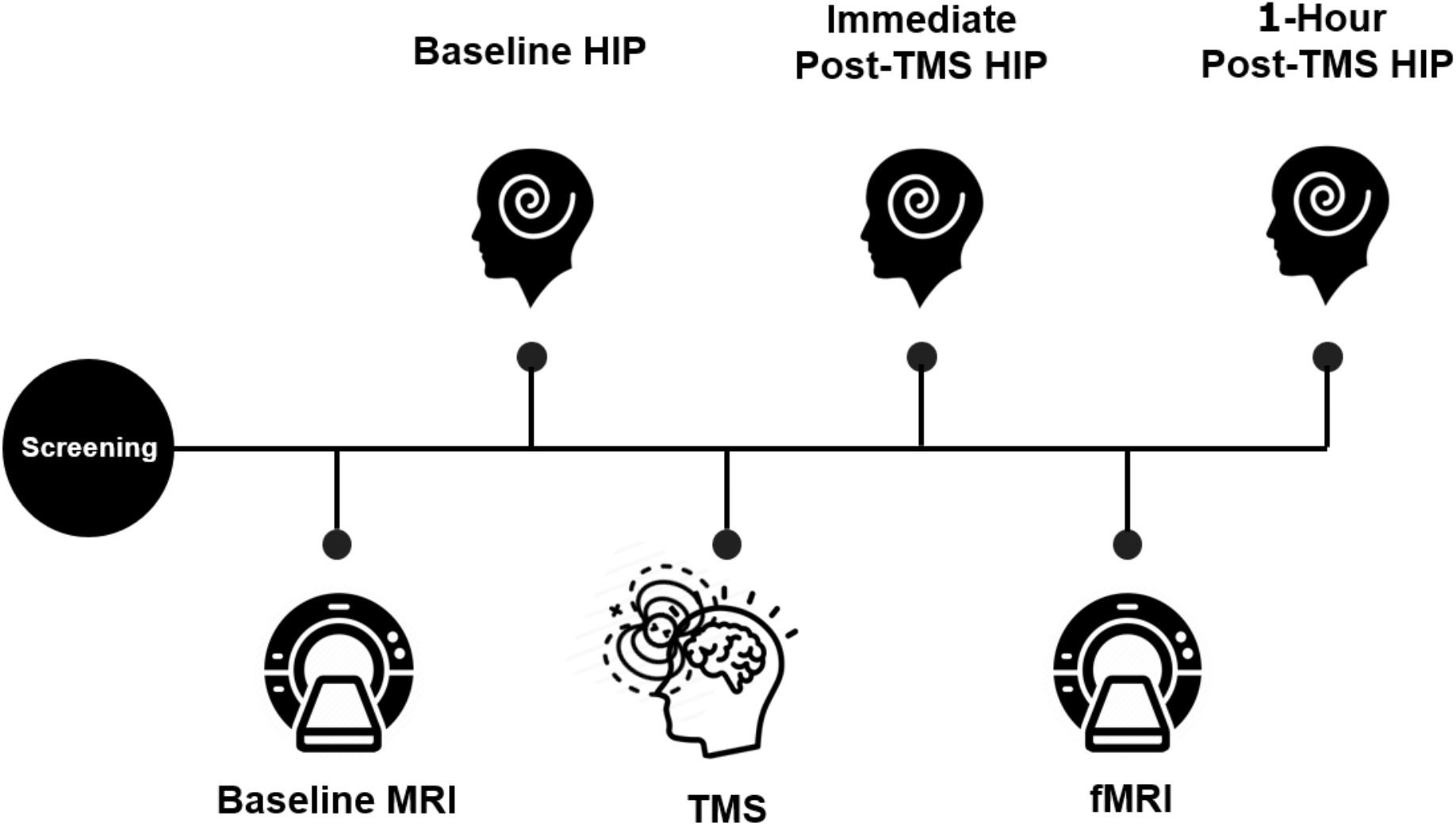
The timeline of study events included the participants undergoing a 1-hour baseline MRI scanning session in which both a structural and functional MRI sequence were completed. These MR images were then used to individually target the cTBS treatment. Participants then received either active or sham cTBS targeted to the L-DLPFC. Hypnotizability was assessed pre, post, and 1-hour post cTBS using the Hypnotic Induction Profile. Abbreviations: HIP = Hypnotic Induction Profile; fMRI = Functional Magnetic Resonance Imaging; TMS = Transcranial Magnetic Stimulation

### Neuroimaging for TMS Targeting

MRI data were collected using a research dedicated 3.0-tesla General Electric Discovery MR750 instrument with a Nova Medical 32-channel head coil. Individualized neuroimaging for subsequent transcranial magnetic stimulation targeting consisted of both structural and functional MRI sequences. A whole-brain 0.9 mm^3^ three-dimensional T1-weighted MPRAGE structural scan was acquired as well as an ∼8-minute resting-state functional MRI scan with the following parameters: simultaneous multi-slice EPI sequence with an acceleration factor of 3 (SMS = 3) with the following parameters: resolution = 1.9mm^3^, TE = 30ms, TR = 1.998s, Flip angle = 77°, slice thickness = 1.8mm, FOV = 230mm.

Prior to the start of resting-state data acquisition, participants were instructed to keep their eyes open for the duration of the scan and look at a fixation cross (black background with white “+” in the center of the field of view). An infrared camera affixed to the head coil was used to monitor compliance and ensure that participants did not fall asleep during the scan.

### fMRI Analysis for TMS Targeting

Personalized L-DLPFC targets were generated for each participant using the resting-state fMRI hierarchical clustering to determine the anatomical location within the L-DLPFC that exhibited the greatest functional connectivity to the dACC. A similar application of this method was previously used to identify cortical targets for a study using theta burst stimulation (TBS), a form of rTMS, for the treatment of depression^29^. All analyses were conducted in each participant’s own brain space (i.e., not warped to a standardized-brain template). Resting-state scans were pre-processed according to typical methods using Statistical Parametric Software Version 12 (SPM12)^30^. In brief, the resting-state scans were motion-corrected, resliced, spatially smoothed with a 3mm Gaussian kernel, detrended using a linear model of the global signal^31^, and band-pass filtered to preserve the typical resting-state frequencies (0.1 Hz – 0.01 Hz).

The co-registered L-DLPFC ROI, consisting of Brodmann Areas 9 and 46, formed the search area for the optimal TMS coil placement. Two separate algorithms were used to determine coil placement. The first algorithm sorted each of the DLPFC and dACC voxels into functional sub-units using a hierarchical agglomerative clustering algorithm (described in detail in supplementary material section). The decision-making algorithm considers the net correlation/anti-correlation amount for each L-DLPFC subunit with all the voxels of the dACC. This value is calculated using the sum of all the correlation coefficients multiplied by all the sizes of the dACC subunits. The decision-making algorithm also considers the size of the L-DLPFC subunit (larger clusters are easier to target) and the spatial concentration of voxels that make up the subunit. The identified L-DLPFC subunit referred to within this manuscript as the target was then loaded into Localite TMS Navigation software (Localite GmbH, Sankt Augustin, Germany) and used to guide the TMS coil placement for each study participant.

### Transcranial Magnetic Stimulation

Participants were randomized using a permuted-block design with varying block sizes to receive either sham or active TMS using a MagVenture MagPro X100 (MagVenture A/S, Farum, Denmark) system with a Cool-B65 A/P coil. Each participant was assigned a numeric active/sham code by an independent study consultant for the duration of the study. The code was then entered into the TMS system before treatment to blind both study staff and participants. For all participants, sham electrodes were placed under the TMS coil adjacent to the hairline. Sham consisted of electrical stimulation built into the coil at a setting of 7/10 (“Sham 7”; *N* = 16) or 1/10 (“Sham 1”; *N* = 22), with “Sham 7” being a higher intensity electrical stimulation. Electrical stimulation in and of itself may result in neuromodulation^32,33^. Thus, low and high sham settings were implemented to control for possible confounding effects of different sham settings. Active stimulation consisted of an inhibitory stimulation protocol applied to the L-DLPFC during the one-hour gap between pre-/post-TMS MRI sessions. Continuous Theta Burst Stimulation (cTBS) is a form of rTMS that has been shown to produce fewer adverse side effects, enhanced outcomes, and longer-lasting treatment effects^34,35^. The spatial location of the L-DLPFC stimulation was identified for each subject independently by analyzing their baseline resting-state fMRI data described in the section above. The stimulation parameters consisted of two ∼46-second applications of cTBS comprised of 800 pulses, 200 pulses for ramping up slowly with 600 pulses at full intensity, delivered in a continuous train with each burst containing 3 pulses at 30Hz repeated at 6Hz^36,37^. To ensure the inhibitory effects of the stimulation persisted through the duration of the 1-hour MRI scan protocol and to achieve maximal neuronal depression, the two applications of inhibitory stimulation were spaced 15-minutes apart^37–42^, at 80% resting motor threshold^42,43^ adjusted to the depth of target (described below). Depth adjustments were performed in cases where the L-DLPFC target-to-scalp distance (*Dt*) was greater than that of the motor hand knob-to-scalp *(Dm*). The L-DLPFC target center of gravity and the motor hand knob vertex were both visually identified using MagVenture’s Localite Neuronavigation software. The respective depths of each site were calculated with a standard automated function within the Localite toolbox. Depth-adjusted stimulation intensity was then calculated with a distance-effect gradient *(g)* of 3% and administered in accordance with the depth adjustment formula discussed by Stokes and colleagues^44–46^. Adjusted treatment intensity (Tx) was prohibited from reaching > 120% resting motor threshold (rMT, measured from the right abductor pollicis brevis muscle).

Tx (*% MSO)* = 0.8 × ([rMT] + *3*(*Dt* − *Dm*)) MSO=maximum stimulator output

Following cTBS application, participants were asked to refrain from discussing information pertaining to the stimulation with study personnel, including those conducting HIP assessments. As an additional blinding measure, HIP assessors did not perform the cTBS procedure and were not present in the room during stimulation. Study personnel (not conducting behavioral assessments) administered a questionnaire to assess participant blinding, which included a binary question of whether they thought they received active or sham stimulation.

### Hypnotic Induction Profile

The Hypnotic Induction Profile (HIP) is a common validated measure of hypnotizability^2,47,48^. The HIP includes a standardized hypnotic induction followed by a set of specific suggestions. The HIP is scored by the administering clinician based on behavioral responsiveness and reports of the examinee’s subjective experience. HIP scores range from 0 (no responsiveness) to 10 (most responsive), with scores above 8 representing highly hypnotizable individuals. In the current study, the HIP was first administered immediately prior to L-DLPFC stimulation, immediately following the cTBS stimulation, and again after the MRI, approximately 1-hour following cTBS.

### Data Analysis

To test the change in HIP scores following cTBS, pre- to post-cTBS changes in HIP scores (i.e., *Δ*HIP) were calculated by subtracting immediate post-from pre-cTBS scores. Dienes and Hutton^25^ showed an averaged change in responsivenesss to hypnotic suggestions of 6% following TMS to L-DLPFC vs. acontrol site. Here, to mitigate the possibility of confounds, we attempted to exclude outliers who’s chang in hypnotizability after the stimulation might have ben impacted by reasons other than the stimulatiion itself. To minimize bias, outlier analysis was done on the entire sample, regardless of group assignment. Outliers were conservatively defined as having *Δ*HIP scores ≥ ±3 standard deviations from the mean (a change of approximately 5 points or more of the 0-10 HIP scale; i.e., 50% percent change in hypnotizability), which resulted in two participants being excluded. Two-tailed t-tests were used for group comparisons of *Δ*HIP scores between the Active and Sham groups. To test whether the immediate post-cTBS mean HIP, adjusted for pre-cTBS scores, differed between the Active and Sham groups, we used an ANCOVA on immediate post-cTBS HIP scores, with group as independent variable and pre-cTBS scores as a covariate. Within-group pre-post changes in HIP scores were tested using one-sample t-tests on *Δ*HIP. We examined the sensitivity of our conclusions to deviation from normality using the nonparametric Mann-Whitney U test and Wilcoxon signed-rank test.

### Power Estimation

Power analysis was conducted focusing on our primary comparison between the cTBS and sham groups in terms of the change in HIP score (*Δ*HIP), which is our primary outcome (significance level of .05, two-tailed). According to Dienes and Hutton^25^, the differential effect of cTBS over the L-DLPFC yielded a medium effect (Cohen’s *d* = .6) on responsiveness to hypnotic suggestions. Given a Cohen’s *d* of .6, 72 to 90 participants is needed (respectively) for the independent t-tests for the primary group comparison, and of 54 to 68 participants (respectively) for the one-sample t-tests for within group comparison between pre and post treatment assessments. In the current study, 78 participants were included in the analyses, indicating adequate power to identify medium effects.

## RESULTS

Based on the intention to treat comparison, *Δ*HIP scores were greater in the Active cTBS group (*M* = .63±1.18) compared to the Sham group (*M* = .01±1.02). This difference was statistically significant (*t(76)* = 2.472, *p* = .016; Figure 3) and yielded a medium effect size (Cohen’s *d* = .56). Within-group t-tests indicated that while the Active group had a statistically significant change in HIP scores from pre- and immediate post-cTBS (*t(39)* = 3.344, *p* = .002; medium effect size: Cohen’s *d* = .53), the Sham group did not show a significant difference (*t(37)* = .040, *p* = .968; Cohen’s *d* = .01). Furthermore, the ANCOVA model (*F(2,75) =* 307.930, *p* < .001) indicated a significant treatment effect (Active vs. Sham; *F(1,75)* = 4.908, *p =* .030; partial *η*^*2*^ = .061) after adjusting for baseline pre-cTBS HIP scores (*F(1,75)* = 612.952, *p <* .*001*). The adjusted immediate post-cTBS HIP mean scores were 6.001±.174 for the Active group and 5.446±.179 for the Sham group (adjusted mean difference = .556 [.056, 1.055]). The data met the assumption for homogeneity of variances.

**Figure 3:**
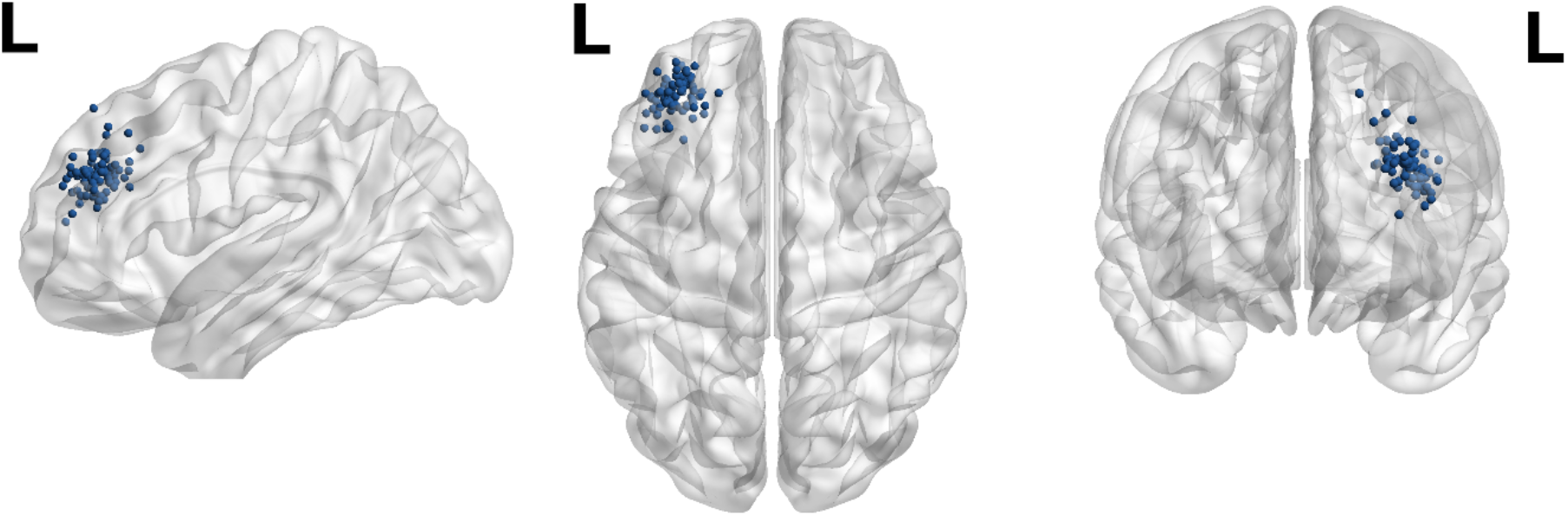
Personalized L-DLPFC neurostimulation targets (blue) for all participants were used in comparison to commonly used beamF3 skull-based measurement coordinates (−35.5, 49.4, 32.4). Although shown here in Montreal Neurological Institute (MNI) standard space for illustration purposes, individual targets were analyzed and identified in native subject space representing the greatest L-DLPFC-dACC functional connectivity.

### Time effects

When tested again approximately 1-hour post-cTBS, the pre- to post-cTBS difference in HIP scores was still significant in the Active group (*Δ*M = .48±1.15, *t(39)* = 2.638, *p* = .012, medium-small effect size: Cohen’s *d* = .42) and not in the Sham group (*Δ*M = .20±1.35, *t(36)*= .912, *p* = .368, Cohen’s *d* = .15; one participant was missing 1-hour post HIP scores). Although 1-hour *Δ*HIP scores were greater in the Active cTBS group compared to the Sham group, the difference between the groups at 1 hour was not statistically significant (*t(75)* = .975, *p* = .333, Cohen’s *d* = .22; See Figure 3). Additionally, after adjusting for baseline pre-cTBS HIP scores, ANCOVA analysis (Corrected Model; *F(2,74) =* 257.197, *p* < .001) indicated that treatment effects were not significant 1-hour post stimulation (Active vs. Sham; *F(1,74)* = .898, *p =* .346; partial *η*^*2*^ = .012), suggesting that the effects of cTBS on hypnotizability might dissipate over time. Adjusted 1-hour post-cTBS HIP mean scores were 5.830±.200 for the Active group and 5.555±.208 for the Sham group (adjusted mean difference = .275 [-.304, .854]).

### cTBS Sham Modes

We tested whether the sham cTBS setting (sham setting 1/10 vs. sham setting 7/10) impacted the difference between the Active and Sham groups. Immediate *Δ*HIP scores were significantly different between the Active and Sham 1 groups (*Δ*M = .81, *t(60)* = 2.821, *p* = .006; Figure 4) with a large effect size (*d* = .78). Conversely, *Δ*HIP scores were not significantly different between the Active and Sham 7 groups (*Δ*M = .36, *t(54)* = 1.026, *p* = .309, *d* = 0.30), indicating that Sham 7 may have had a small neuromodulatory effect. Sham 1 and Sham 7 *Δ*HIP scores were not significantly different (*Δ*M = -.45, *t(36)* = -1.356, *p* = .184). No association was found between sham TMS setting and participants’ guesses as to whether they received Active or Sham stimulation (*X*^2^(2) = 1.878, *p* = .391), indicating that the blind was maintained throughout active, sham 7, and sham 1 settings.

**Figure 4:**
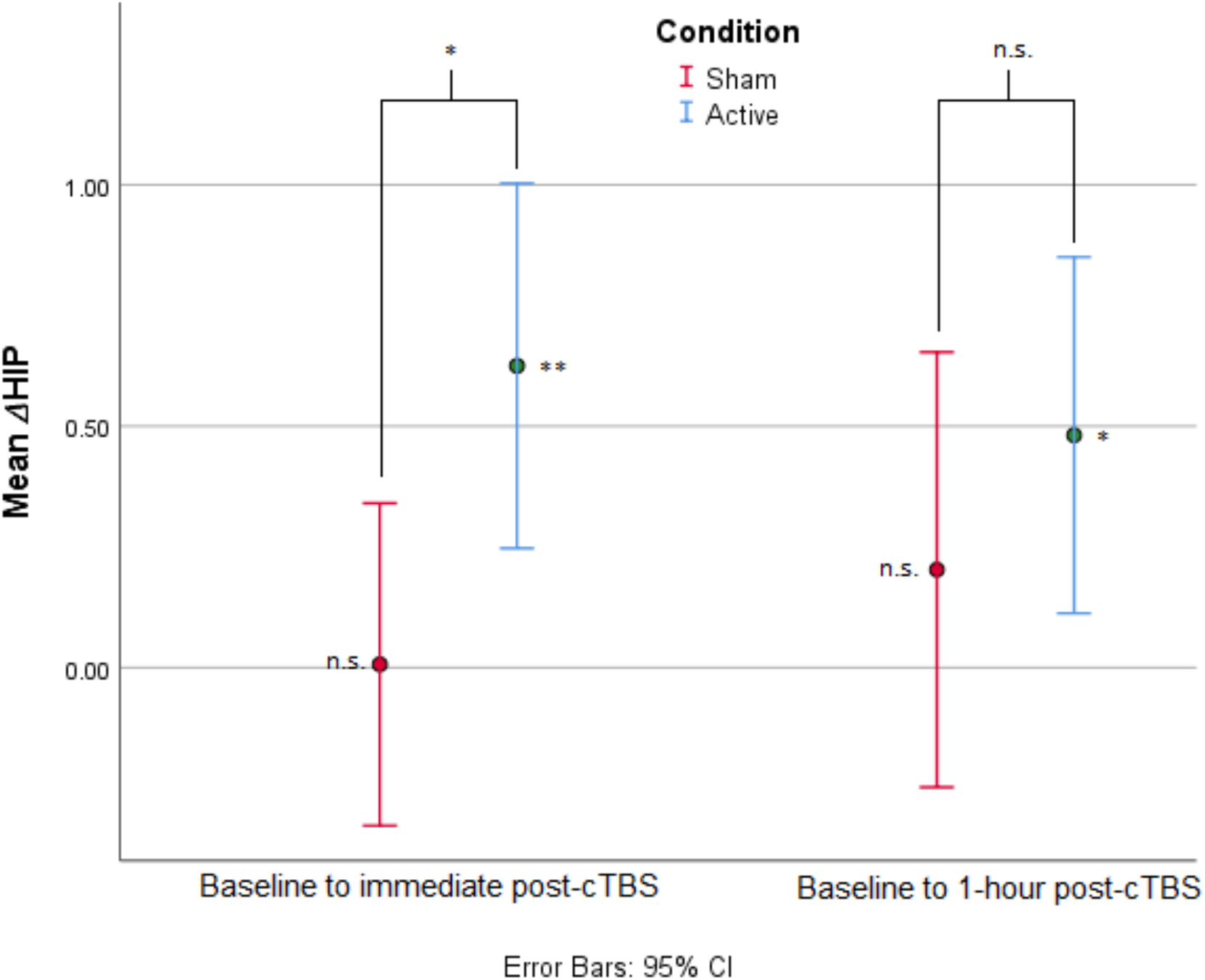
Mean ΔHIP scores comparing pre-cTBS to (a) immediately post-cTBS, and (b) 1-hour post-cTBS in the Active (in blue) and Sham (in red) groups. Immediate ΔHIP scores represented a significant difference from baseline in the Active cTBS group but not the Sham group. Similarly, 1-hour ΔHIP scores represented a significant difference from baseline in the Active cTBS group but not the Sham group. ΔHIP scores were significantly greater in the Active cTBS group than in the Sham group immediately post-cTBS, but not 1-hour post-cTBS. * p < .05; ** p < .01

**Figure 5:**
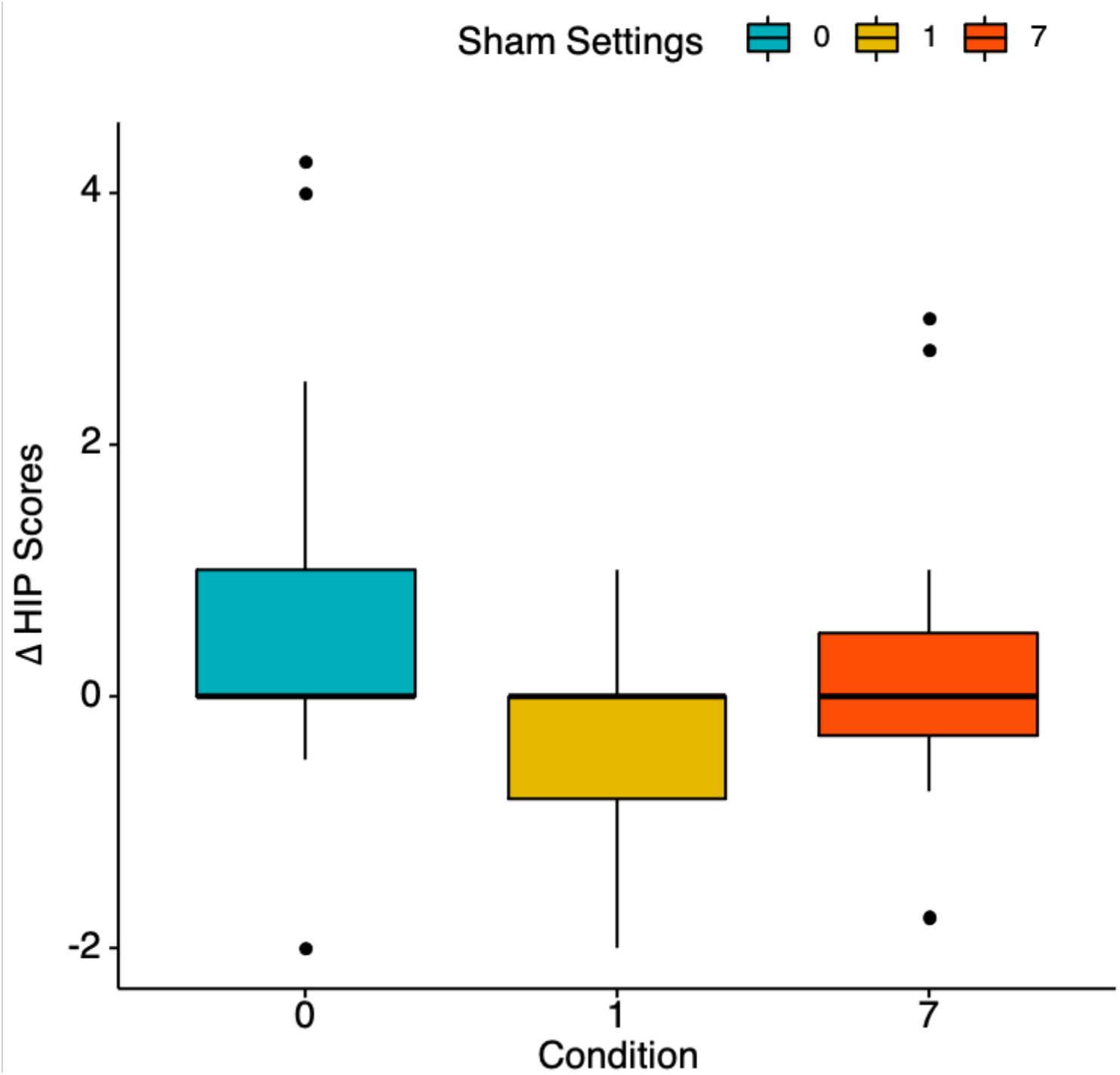
Bar graph of mean ΔHIP scores comparing Active (in blue) and the two Sham cTBS settings: 1/10 (Sham 1; in yellow) and 7/10 (Sham 7; in red). ΔHIP scores were significantly different between Active and Sham 1, but not between Active and Sham 7. Sham 1 and Sham 7 ΔHIP scores were not significantly different from one another.

## DISCUSSION

Using noninvasive neurostimulation, we demonstrated immediate modulation of a stable, clinically relevant neurobehavioral trait. Building on our group’s previous work on the neural bases of hypnotizability^4,20,49^, our results suggest a causal relationship between L-DLPFC inhibition and hypnotizability, which was previously shown to be associated with DLPFC-dACC functional connectivity in a healthy population^4^. Since we have also found that entering a state of hypnosis is associated with reduced activity in the dACC^49^, our findings further support the hypothesis that DLPFC-dACC connectivity is a salient neurocognitive component of hypnotizability. Further investigations of neuroimaging correlates are needed to confirm pre-post stimulation changes in DLPFC-dACC connectivity and their relationship with changes in hypnotizability.

We utilized cTBS, a neuromodulation approach that has been both documented as safe^43^ and effective at facilitating functional connectivity^24^. Specifically, two cTBS sessions, spaced 15-minutes apart, have been shown to elicit effects lasting >1 hour, enabling a window of time augment hypnotizaiblty with particular relevance to application in a clincal setting^37,50^. cTBS approaches that mimic intrinsic physiological and temporal rhythms may induce synaptic plasticity analogous to long-term potentiation (LTP) and/or long-term depression (LTD) as demonstrated in animal models^51^, providing a cellular basis for changes in behavioral phenotype.

We utilized a conservative approach with our cTBS treatment parameters in that we aimed to modulate the targeted network only long enough to allow for our neuroimaging sequences which required 1 hour. This conservative approach achieved medium to large effect sizes, yet the mean increase in hypnotizability scores was relatively small. In the longitudinal study conducted by Piccione et al.^7^, it was found that trait hypnotizability increased only 5% on average across 25 years. Indeed the test-retest correlation was .7 over that time interval. Here, we demonstrated an increase of 6% on average in hypnotizability following 92 seconds of noninvasive neurostimulation. Providing evidence of a mechanistic pathway for TMS to modulate stable traits is encouraging, as the intensity of clinical effects can be adjusted by the specific stimulation regimen. For example, building on the evidence of mechanistic pathways for TMS to modulate depression, we recently demonstrated that conventional rTMS treatment of depression could be condensed and improved substantially by increasing the pulse potency and shortening intersession intervals^29^. Similar optimization of dose-response should be further examined to identify whether a different stimulation protocol can lead to a more clinically beneficial increase in hypnotizability (i.e., one that would render hypnosis-based treatment significantly more effective) by the neural pathway we portrayed.

This study serves as proof-of-concept of the possibility of modulating a neural trait in patients with FMS. This is particularly encouraging, as functional connectivity in the default mode network (DMN) and the salience network, both involved in hypnosis^49^ and targeted in this study, have been shown to be altered in persons with FMS^52^. Moreover, other chronic pain populations have also shown differences in functional connectivity in the salience network^53,54^. As our group has previously shown, persons who are innately more highly hypnotizable show greater functional connectivity between the DLPFC and the dACC^4^ and conditions that influence connectivity, such as fibromyalgia, may influence the degree to which the trait may be modulated. Even though we showed that this trait was able to be modulated with cTBS in this population, it is possible that lower baseline connectivity in our FMS sample renders the enhancement of hypnotizability through this route rather challenging. Alternatively, depending on the underlying pathophysiology of the condition, specific neuromodulation paradigms may be warranted to enhance hypnotizability without reliance on this pathway.

As TMS is limited by target depth, we indirectly modulated the dACC through its established functional connectivity to the DLPFC. Therefore, we targeted the region of the L-DLPFC with the highest functional connectivity to the dACC. We demonstrated that this neuromodulatory technique is feasible, effective, and safe; however, we recognize that alternative forms of noninvasive neuromodulation may be able to produce similar or even more efficacious results. Specifically, neuromodulation of deep cortical or subcortical structures through techniques that are not limited by tissue penetration constraints such as low-intensity focused ultrasound, would enable an alternative approach to directly modulate the dACC and investigate reciprocal DLPFC-dACC connectivity. Further, direct modulation of neural activity in either the DLPFC or the dACC would provide more definitive evidence of the importance of each region in hypnotizability.

Alongside our novel evidence for the feasibility of modulating a stable neural trait through neuromodulation, previous research has demonstrated the modulation of more transient traits using TMS. For example, Möbius et al.^55^ has modulated the susceptibility to mood induction using excitatory rTMS (10 Hz). Spronk et al.^56^ observed a significant decrease in trait neuroticism and an increase in extraversion following ten rTMS sessions applied to the L-DLPFC. The modulation of trait neuroticism was later replicated by Berlim et al.^57^. This is notable as, beyond time-dependent changes in trait neuroticism, treatment for depression largely fails to modulate it^58^. Taken together with our findings, rTMS may be able to modulate clinically relevant neural traits associated with psychopathology and responsiveness to treatment.

Furthermore, we observed potential qualitative differences between sham intensity settings. Differences between sham settings could be explained through two routes, differential effects of electrical stimulaion or placebo. E-field modeling by Smith and Peterchev^32^ demonstrated that higher active TMS intensities would result in proportionately higher sham E-field strengths. Therefore, using a higher sham stimulation setting likewise increases the risk of inducing unintentional neural effects. While the effects of different sham stimulations on neural networks need to be further investigated, a placebo effect might be involved if those sham stimulations are experienced differently on the scalp. Studies examining whether chronic pain populations, including FMS, are more responsive to placebo than non-pain healthy controls show mixed findings^59–61^; however, a systematic review investigating placebo-intervention against no-treatment comparator groups in FMS found that the magnitude of placebo effects increases with the effect size of the active treatment^62^. While it is possible that a higher intensity sham stimulation would give rise to a stronger placebo response, we observed no significant differences in participants’ guesses as to whether they received sham or active treatment between either sham group nor the active group. Therefore, it is unlikely that the placebo response, if involved, differed substantially between the two sham settings we used. Based on our observation that a lower sham setting yielded a rather similar blind as the higher setting, we suggest adjusting the sham settings to 1/10, as we identified a smaller active vs. sham effect using greater sham stimulation levels.

## Limitations

The interpretation of the current results should consider several caveats that could be addressed in future trials. As aforementioned, we utilized a conservative stimulation approach in this initial trial to limit the modulation effects to only 1 hour. Future studies may observe larger effect sizes by increasing the number of sessions or total pulse dose administered to patients. Additionally, our study assessed the effects of two different sham settings to assess the result that a higher sham setting would have on the post-stimulation outcome measure. While these two settings showed a difference, our study was not designed to determine if this difference was due to a weak neuromodulatory effect or if the increased sham sensation contributed to an increase in the placebo effect. Finally, this trial did not assess any clinical outcome measures as this was designed to be a mechanistic study; future studies should build upon these findings to assess the use of neuromodulation of a neural trait to directly assess clinical outcome measures in a patient population.

The results achieved in this study provide evidence that stable neural traits can be predictably, directionally, and measurably modulated. Further studies are needed to build upon these findings to better understand the individual elements driving this change in a neural trait and how neuromodulation can be integrated into best clinical practices for treating patients with trait-based disorders as well as enhancing trait-based interventions.

## Data Availability

The data that support the findings of this study are available from the corresponding authors, NW and DS, upon reasonable request.

## ACKNOWLEDGEMENTS

This work was supported by the National Institute of Health (NIH) Center for Complementary and Integrative Health (NCCIH) grant Innovation Award for Mechanistic Studies to Optimize Mind and Body Interventions (R33AT009305-03; D.S. & N.R.W.). The authors would like to thank Keith Sudheimer for his technical assistance.

